# Coronavirus Disease (COVID-19) Pandemic: An Overview of Systematic Reviews

**DOI:** 10.1101/2020.04.16.20068213

**Authors:** Israel Júnior Borges do Nascimento, Dónal P. O’Mathúna, Thilo Caspar von Groote, Hebatullah Mohamed Abdulazeem, Ishanka Weerasekara, Ana Marusic, Livia Puljak, Vinicius Tassoni Civile, Irena Zakarija-Grkovic, Tina Poklepovic Pericic, Alvaro Nagib Atallah, Santino Filoso, Nicola Luigi Bragazzi, Milena Soriano Marcolino, On behalf of the International Network of Coronavirus Disease 2019 (InterNetCOVID-19)

**Author notes:** Corresponding author: Israel Júnior Borges do Nascimento, ClinPath, University Hospital and School of Medicine, Universidade Federal de Minas Gerais, or.

## Abstract

**Background:** Navigating the rapidly growing body of scientific literature on the SARS-CoV-2 pandemic is challenging and ongoing critical appraisal of this output is essential. We aimed to collate and summarize all published systematic reviews on the coronavirus disease (COVID-19).

**Methods:** Nine databases (Medline, EMBASE, Cochrane Library, CINAHL, Web of Sciences, PDQ-Evidence, WHO’s Global Research, LILACS and Epistemonikos) were searched from December 1, 2019 to March 24, 2020. Systematic reviews analyzing primary studies of COVID-19 were included. Two authors independently undertook screening, selection, extraction (data on clinical symptoms, prevalence, pharmacological and non-pharmacological interventions, diagnostic test assessment, laboratory and radiological findings) and quality assessment (AMSTAR 2). Meta-analysis on prevalence of clinical outcomes was performed.

**Results:** Eighteen systematic reviews were included; one was empty. Using AMSTAR 2, confidence in the results of 13 reviews was rated as “critically low”, one as “low”, one as “moderate” and two as “high”. Symptoms of COVID-19 were (range values of point estimates): fever (82–95%), cough with or without sputum (58–72%), dyspnea (26–59%), myalgia or muscle fatigue (29–51%), sore throat (10–13%), headache (8– 12%) and gastrointestinal complaints (5–9%). Severe symptoms were more common in men. Elevated C-reactive protein (associated with lymphocytopenia) and lactate dehydrogenase, and slightly elevated aspartate and alanine aminotransferase, were commonly described. Thrombocytopenia and elevated levels of procalcitonin and cardiac troponin I were associated with severe disease. Chest imaging described a frequent pattern of uni- or bilateral multilobar ground-glass opacity. Only one review investigated the impact of medication (chloroquine) but found no verifiable clinical data. All-cause mortality ranged from 0.3% to 14%.

**Conclusions:** Confidence in the results of most reviews was “critically low”. Future studies and systematic reviews should adhere to established methodologies. The majority of included systematic reviews were hampered by imprecise search strategy and no previous protocol submission.

**Protocol registration:** This is an extension of a PROSPERO protocol (CRD42020170623); protocol available on Open Science Framework (https://osf.io/6xtyw).

## 1. Background

Spread of severe acute respiratory coronavirus 2 (SARS-CoV-2), the causal agent of COVID-19, has been characterized as a pandemic by the World Health Organization (WHO) and has triggered an international public health emergency [1]. The virus is easily transmitted through person-to-person contact and virus contamination of surfaces or objects [2,3]. As of April 7, 2020, more than 1,359,398 people have been confirmed to be infected with the virus and over 75,945 people have died as a result of severe complications of the disease [4]. These numbers are likely to be underestimates, as countries are unable to test all patients suspected of having the disease due to worldwide shortages of testing reagents [5]. Estimates put the economic impact of COVID-19 as surpassing trillions of dollars when global trade, travel and business costs are added to healthcare and public health expenses [6,7].

Given the novelty of the situation and dozens of daily publications in peer reviewed journals and preprints, there is an overwhelming avalanche of information from research articles to sift through. For healthcare professionals and government policy makers, the ability to sort through relevant research results, in order to clearly identify the most up-to-date and reliable information, is essential. Having filtered, accurate information at their disposal allows stakeholders to make timely decisions regarding the best course of action. Hence, our aim was to summarize the data from all available systematic reviews regarding COVID-19 in humans and to rigorously assess the level of confidence that we can have in this evidence.

## 2. Methodology

### 2.1. Protocol and Registration

This overview is a systematic review of systematic reviews. It is an extension of a recently submitted PROSPERO protocol (CRD42020170623) [8]. The protocol has also been published on Open Science Framework (OSF) (https://osf.io/6xtyw) for transparency. It was conducted in accordance with the Preferred Reporting Items for Systematic Reviews and Meta-Analyses (PRISMA) statement [9]. The methodology used in this review was adapted from the *Cochrane Handbook for Systematic Reviews of Interventions* and also followed established methodological considerations for analyzing existing systematic reviews [10,11].

### 2.2. Ethics

Approval of a research ethics committee was not necessary as the study analyzed only publicly available articles.

### 2.3. Eligibility Criteria

Systematic reviews were included if they analyzed primary data from patients infected with SARS-CoV-2 as confirmed by RT-PCR or another pre-specified diagnostic technique. Eligible reviews covered all topics related to COVID-19, including, but not limited to those that reported clinical symptoms, diagnostic methods, therapeutic interventions, laboratory findings or radiological results. Both full manuscripts and abbreviated versions, such as letters, were eligible.

No restrictions were imposed on the design of the primary studies included within the systematic reviews, date of last search, whether or not the review included meta-analyses, or language. A systematic review was defined as a publication where: I. the search was conducted in at least two databases (at least one being electronic); and II. detailed description of the methods with explicit inclusion criteria was provided. We excluded narrative reviews as these are less likely to be replicable and are more prone to bias.

### 2.4. Information Sources

Nine databases were searched for eligible records published between December 1 2019 and March 24, 2020: Cochrane Database of Systematic Reviews via Cochrane Library, PubMed EMBASE, CINAHL (Cumulative Index to Nursing and Allied Health Literature), Web of Sciences, LILACS (Latin American and Caribbean Health Sciences Literature), PDQ-Evidence, WHO’s Global Research on Coronavirus Disease (COVID-19), and Epistemonikos.

### 2.5. Search

The comprehensive search strategy for each database is provided in Appendix 1 and was designed and conducted in collaboration with an information specialist. All retrieved records were primarily processed in EndNote, where duplicates were removed, and records were then imported on Covidence Platform [12]. In addition to database searches, we screened reference lists of reviews included, after screening records retrieved via databases.

### 2.6. Study Selection

All searches, screening of titles and abstracts, as well as record selection, were performed independently by two investigators using the Covidence Platform [12]. Articles deemed potentially eligible were retrieved for full-text screening by two independent investigators. Discrepancies in both stages of screening were resolved by consensus. During screening, records published in languages other than English were translated by a native/fluent speaker.

### 2.7. Data Collection Process

We custom designed a data extraction table for the purpose of this study, which was piloted by two authors independently. Data extraction was performed independently by two authors. Conflicts were resolved by consensus or by consulting a third researcher.

### 2.8. Data Items

We extracted the following data: article identification data (authors’ name and journal of publication), range of period associated with the search, number of databases searched, population or settings considered, main results and outcomes observed, and number of participants. From Web of Science (Clarivate Analytics, Philadelphia, PA, USA) we extracted journal rank (quartile) and Journal Impact Factor (JIF).

We categorized the following as primary outcomes: all-cause mortality, need for and length of mechanical ventilation, length of hospitalization (in days), admission to intensive care unit (yes/no) and length of stay in intensive care unit.

The following were categorized as exploratory outcomes: diagnostic methods used for detection of the virus, male to female ratio, clinical symptoms, pharmacological and non-pharmacological interventions, laboratory findings (full blood count, liver enzymes, C-reactive protein, d-dimer, albumin, lipid profile, serum electrolytes, blood vitamin levels, glucose levels, and any other important biomarkers), and radiological findings (using radiography, computed tomography, magnetic resonance imaging or ultrasound).

We also collected data on reporting guidelines and requirements for publication of systematic reviews and meta-analyses from journal websites where included reviews were published

### 2.9. Quality Assessment in Individual Reviews

Two researchers independently assessed the quality of the reviews using the “A MeaSurement Tool to Assess systematic Reviews 2 (AMSTAR 2)”, designed to assess systematic reviews that include both randomized and non-randomized studies. Adherence to each item was rated as follows: yes, no, partial yes, not applicable or unclear. We rated overall confidence in the results of the review as “critically low”, “low”, “moderate” or “high”, according to the AMSTAR 2 guidance based on seven critical domains defined by AMSTAR 2 authors [13]. If a review did not find any studies to include (i.e., it was an “empty review”), we scored each individual item, but we did not conduct summary confidence assessment for such reviews.

One of the included systematic reviews was conducted by some members of this author team. To prevent risk of bias in assessing methodological quality of this particular review, it was assessed independently by two authors who did not co-author it.

### 2.10. Synthesis of Results

For data synthesis, we prepared a table summarizing each systematic review. Graphics illustrating mortality rate and clinical symptoms were created. We then prepared a narrative summary of the methods, findings, study strengths and limitations.

For analysis of prevalence of clinical outcomes, we extracted data on the number of events and total number of patients to perform proportional meta-analysis using RStudio^©^ software, with the “meta” package (version 4.9-6), using the “metaprop” function for reviews that did not perform meta-analysis, excluding case studies because of the absence of variance. For reviews that did not perform meta-analysis, we presented pooled results of proportions with their respective confidence intervals (95%) by the inverse variance method with a random-effects model, using the DerSimonian-Laird estimator for τ^2^. We adjusted data using Freeman-Tukey double arcosen transformation. Confidence intervals were calculated using the Clopper-Pearson method for individual studies. Forest plots were generated showing data from individual systematic reviews, but results were not pooled because some studies were included in multiple reviews. We developed the using RStudio^©^ software, with the “metafor” package (version 2.1-0) and “forest” function.

## 3. Results

Our search retrieved 1,063 publications, of which 175 were duplicates. Most publications were discarded after title and abstract analysis (n = 860), 28 were selected for full text screening and 18 were included in the final analysis (Figure 1) [14-31]. Reference list screening did not retrieve any additional systematic reviews.

**Figure 1.**
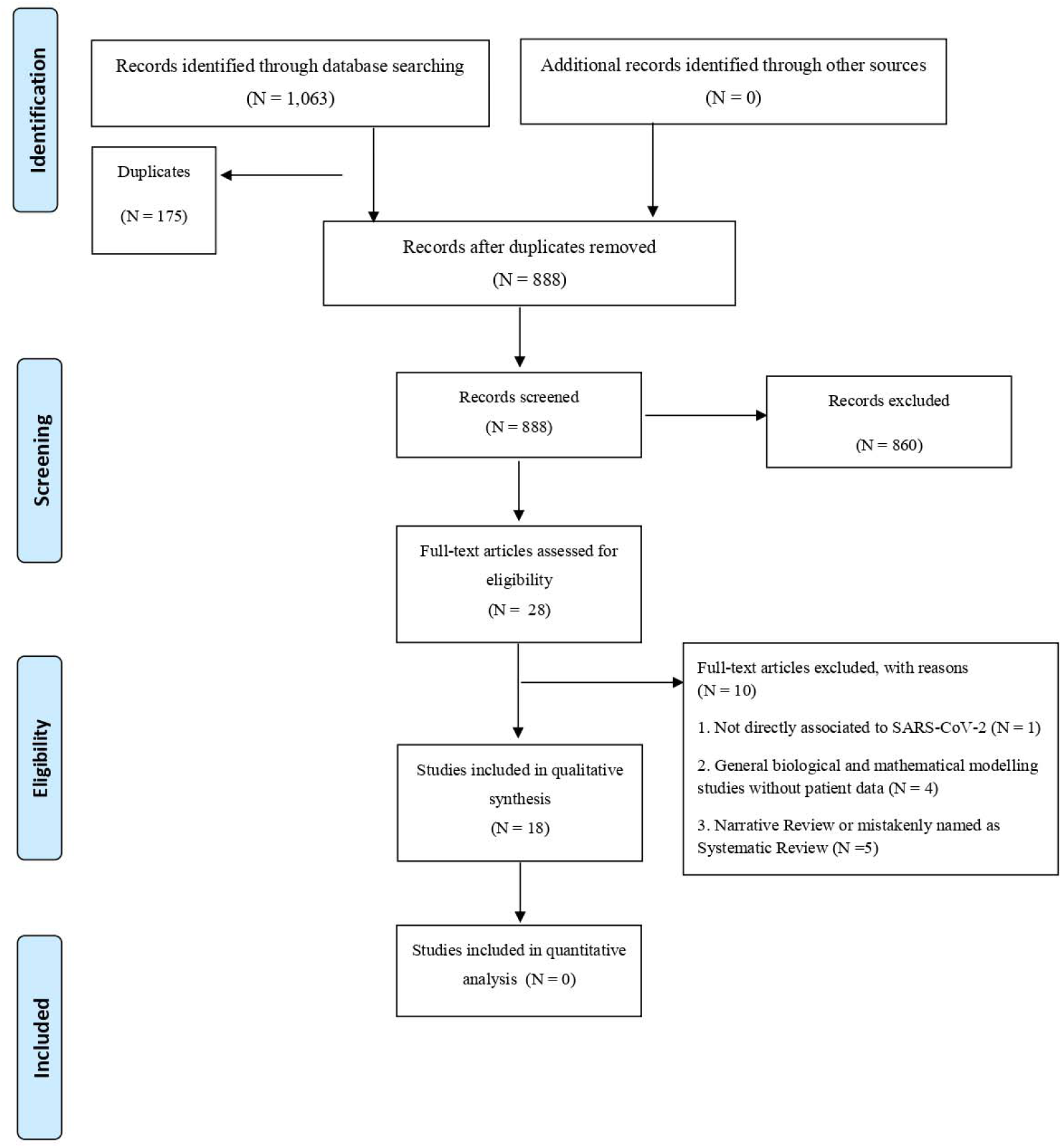
PRISMA flow diagram.

### 3.1. Characteristics of included reviews

Summary features of the 18 systematic reviews are presented in Table S1. Included reviews were published in 14 different journals. Only four of these journals had specific requirements for submission of systematic reviews (with or without meta-analysis): *European Journal of Internal Medicine, Journal of Clinical Medicine, Ultrasound in Obstetrics and Gynecology* and *Clinical Research in Cardiology*. Two journals reported that they published only invited reviews (*Journal of Medical Virology* and *Clinica Chimica Acta*). Three reviews were published as a letter, and one was labelled as a rapid review (Table S2).

All the reviews were published in English in a first quartile (Q1) journal, with JIF (Journal Impact Factor) ranging from 1.692 to 6.062. The 18 systematic reviews covered more than 120 individual primary studies, involving in total at least 60,000 participants. Ten systematic reviews included meta-analyses. Primary studies included in the reviews were published between December 2019 and March 18, 2020, and comprised case reports, case series, cohorts and other observational studies. We did not find any review with randomized clinical trials. In these reviews, systematic literature searches were performed from 2019 (entire year) up to March 9, 2020.

### 3.2. Population and study designs

Most of the reviews analyzed data from patients with COVID-19 who developed pneumonia, acute respiratory distress syndrome (ARDS) or any other correlated complication. Two reviews were based on *in vitro* investigations, one review aimed to evaluate the effectiveness of using surgical masks on preventing transmission of the virus [25], one review was focused on pediatric patients [23] and one review investigated COVID-19 in pregnant women (Figure 1) [26]. Most reviews assessed clinical symptoms, laboratory findings or radiological results.

### 3.3. Systematic review findings

The summary of findings from individual reviews is shown in Table S2. Overall, all-cause mortality ranged from 0.3% to 14% (Figure 2).

**Figure 2.**
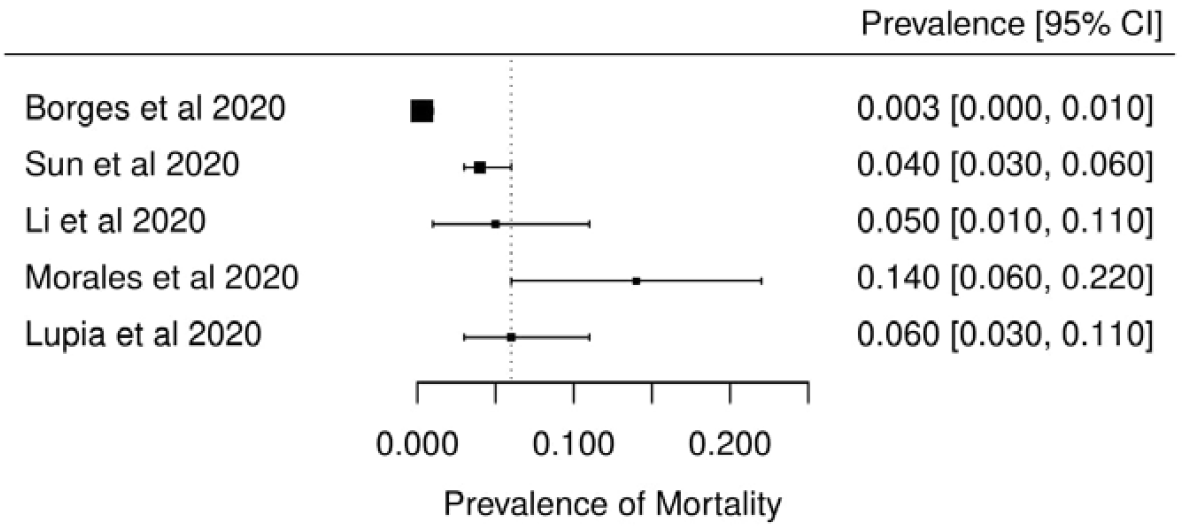
Meta-analysis of prevalence of mortality among five systematic reviews.

#### 3.3.1. Clinical symptoms

Seven reviews described the main clinical manifestations of COVID-19 [14,15,22,23,24,28,30]. Three of them provided only a narrative discussion of symptoms [14,23,24]. Four reviews performed a statistical analysis of the incidence of different clinical symptoms. Overall, symptoms in patients with COVID-19 were (range values of the point estimates): fever (82–95%), cough with or without sputum (58– 72%), dyspnea (26– 59%), myalgia or muscle fatigue (29–51%), sore throat (10–13%), headache (8–12%), gastrointestinal disorders, such as diarrhea, nausea or vomiting (5.0–9.0%), and others (including, in one study only: dizziness 12.1%) (Figure 3-9). Three reviews assessed cough with and without sputum together; only one review assessed sputum production itself (28.5%).

**Figure 3.**
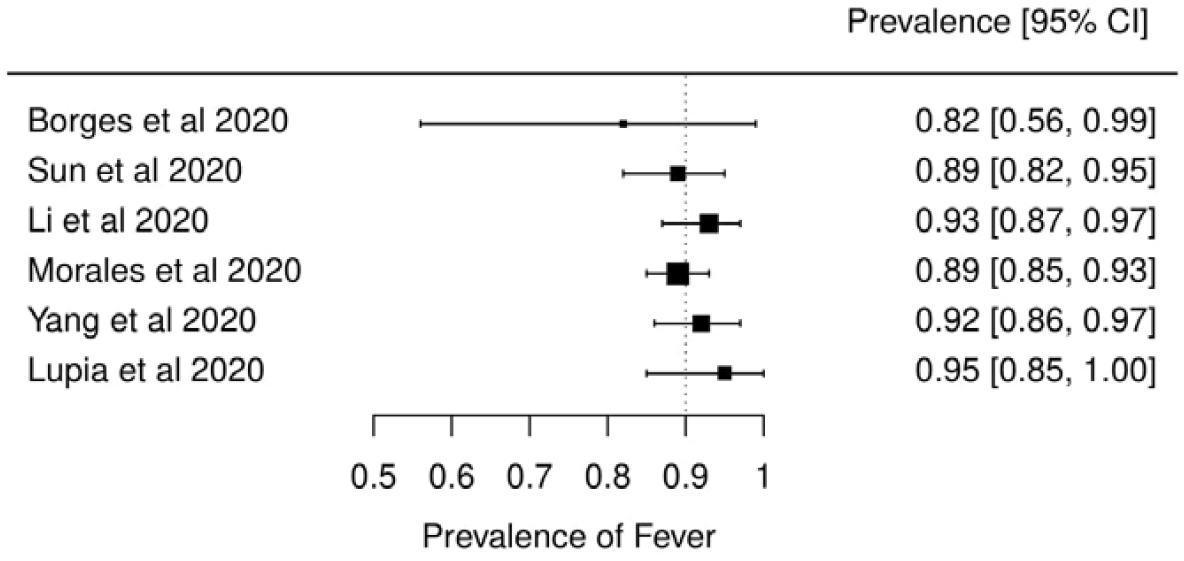
Meta-analysis of prevalence of fever among six systematic reviews.

**Figure 4.**
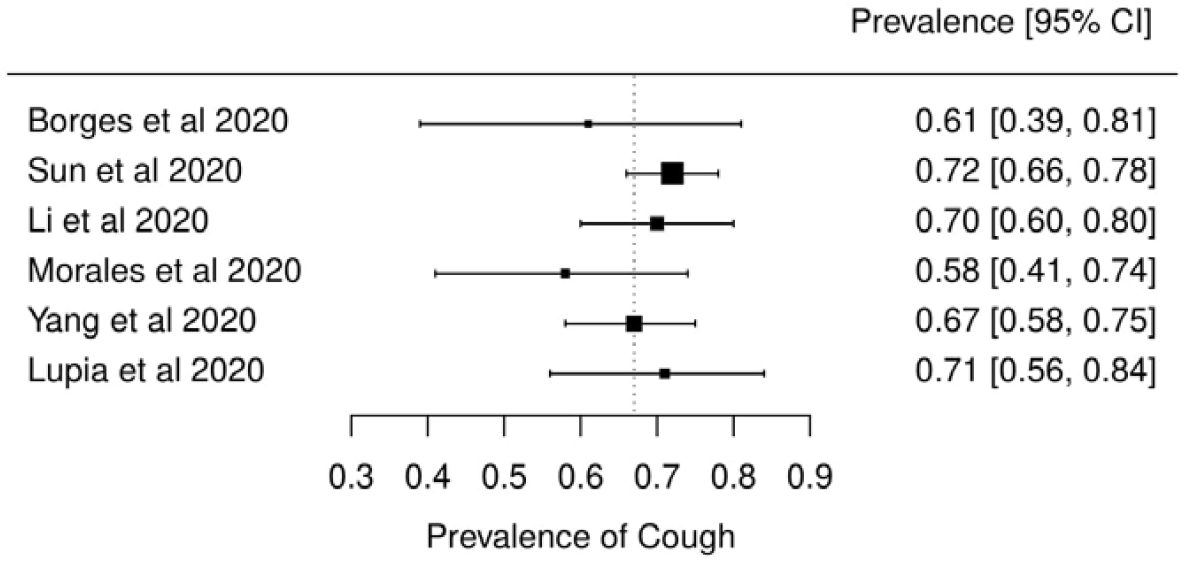
Meta-analysis of prevalence of cough among six systematic reviews.

**Figure 5.**
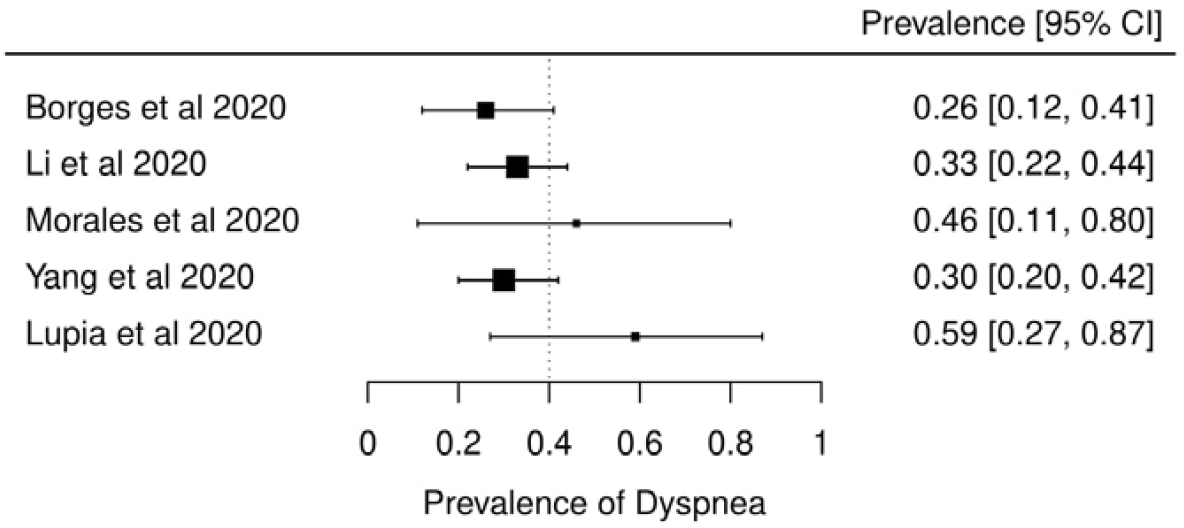
Meta-analysis of prevalence of dyspnea among five systematic reviews.

**Figure 6.**
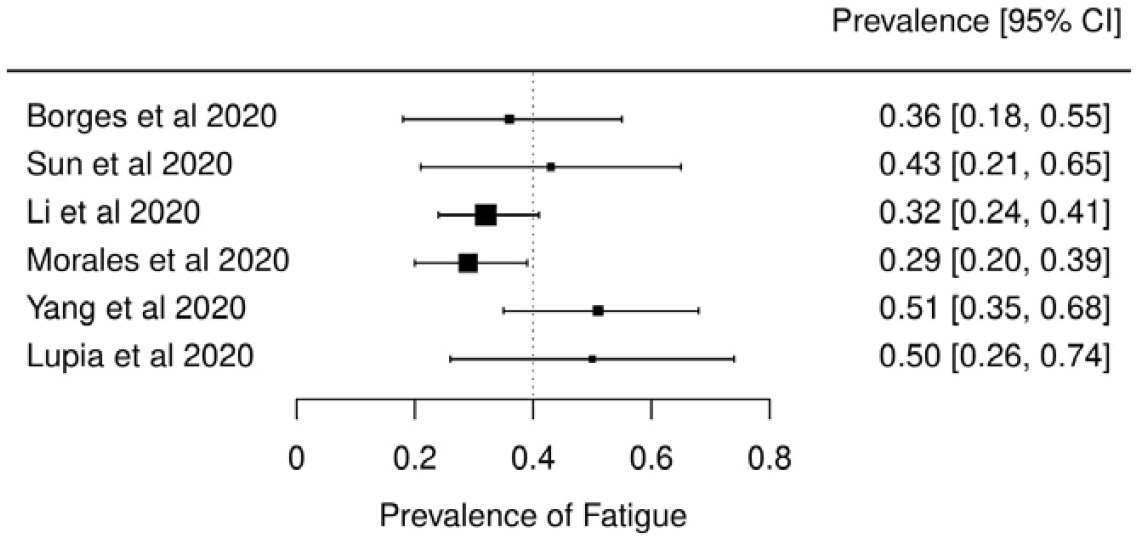
Meta-analysis of prevalence of fatigue or myalgia among five systematic reviews.

**Figure 7.**
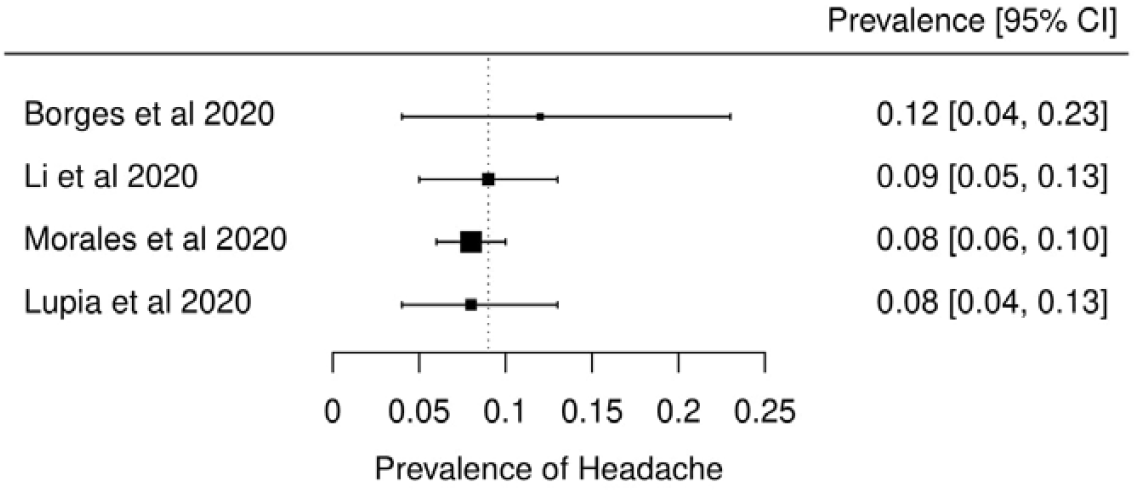
Meta-analysis of prevalence of headache among four systematic reviews.

**Figure 8.**
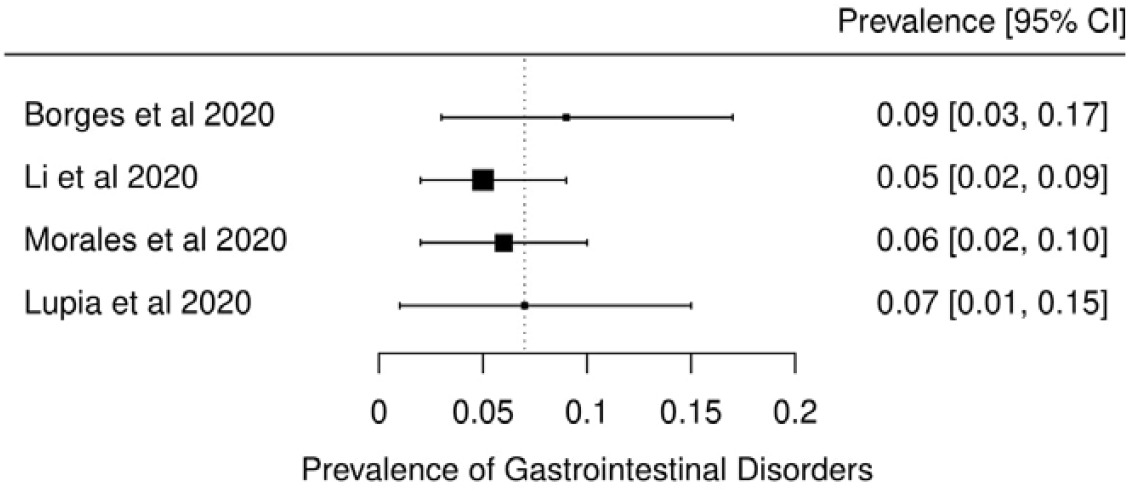
Meta-analysis of prevalence of gastrointestinal disorders among four systematic review

**Figure 9.**
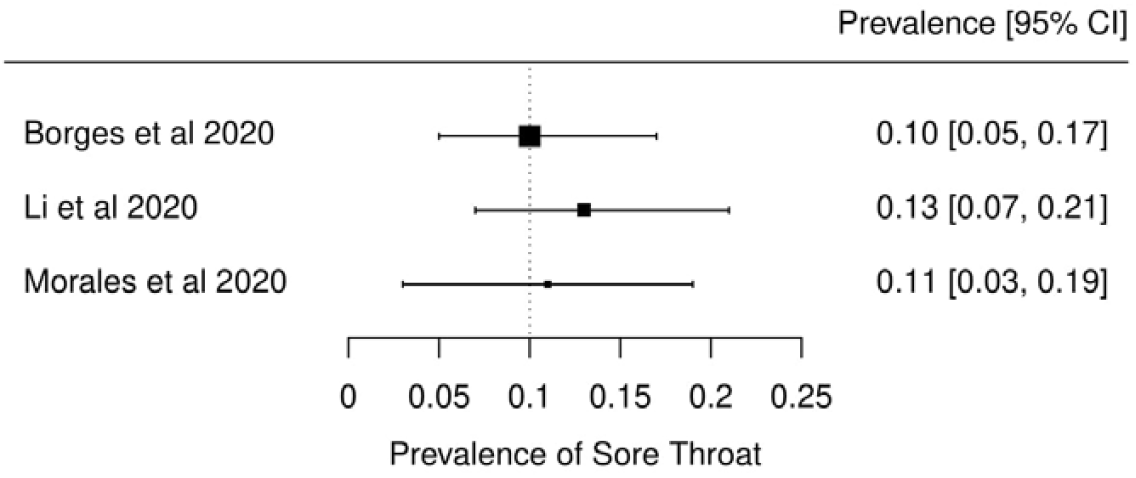
Meta-analysis of prevalence of sore throat among three systematic reviews.

#### 3.3.2. Diagnostic aspects

Three reviews described methodologies, protocols and tools used for establishing the diagnosis of COVID-19 [14,23,27]. Use of respiratory swabs (nasal or pharyngeal) or blood specimens to assess the presence of SARS-CoV-2 nucleic acid using RT-PCR assays was the most commonly used diagnostic methods mentioned in the included studies. These diagnostic tests are widely used, but their precise sensitivity and specificity remain unknown. One review included a Chinese study in which there was clinical diagnosis with no confirmation of SARS-CoV-2 infection (patients were diagnosed with COVID-19 if they presented with at least 2 symptoms suggestive of COVID-19, as well as laboratory and chest radiography abnormalities) [23].

#### 3.3.3. Therapeutic possibilities

Pharmacological and non-pharmacological interventions (supportive therapies) used in the treatment of patients with COVID-19 were reported in five reviews [16,17,23,24,27]. Most reviews were descriptive, as the original studies did not assess the impact of therapies. Antivirals have been used empirically for COVID-19 treatment with the most commonly used being protease inhibitors (lopinavir, ritonavir, darunavir), nucleoside reverse transcriptase inhibitor (tenofovir), nucleotide analog (remdesivir, galidesivir, ganciclovir), and neuraminidase inhibitors (oseltamivir). Umifenovir, a membrane fusion inhibitor, was investigated in several studies [16,24]. Possible supportive interventions analyzed were different types of oxygen supplementation and breathing support (invasive or non-invasive ventilation). Use of antibiotics, both empirically and to treat secondary pneumonia, was reported in some studies. One review specifically assessed evidence on the efficacy and safety of the anti-malaria drug chloroquine [17]. It identified 23 ongoing trials investigating the potential of chloroquine as a therapeutic option for COVID-19, but no verifiable clinical outcomes data. Use of mesenchymal stem cells, antifungals and glucocorticoids were seldom described.

#### 3.3.4. Laboratory and radiological findings

Of the 18 reviews included in this overview, eight analyzed laboratory parameters in patients with COVID-19 [16,19-24,28]. Elevated C-reactive protein levels, associated with lymphocytopenia, elevated lactate dehydrogenase, as well as slightly elevated aspartate and alanine aminotransferase (AST, ALT) were commonly described. Lippi et al. assessed the variation of cardiac troponin I (cTnI), procalcitonin, and platelet count between severe COVID-19 patients and non-severe COVID-19 patients [19-21]. Elevated levels of procalcitonin and cTnI were more likely to be associated with a severe disease course (requiring intensive care unit admission and intubation). Furthermore, thrombocytopenia was frequently observed in patients with complicated COVID-19 infections.

Chest imaging (chest radiography and/or computed tomography) features were assessed in six reviews, all of which described a frequent pattern of local or bilateral multilobar ground-glass opacity [16,23,24,28-30]. Septal thickening, bronchiectasis, pleural and cardiac effusions, halo signs and pneumothorax were observed in patients suffering from COVID-19.

#### 3.4 Quality of evidence in individual systematic reviews

Table S3 shows the detailed results of the quality assessment of 18 systematic reviews, including assessment of individual items and summary assessment. Overall confidence in results of reviews was assessed for 17 reviews; for one empty review summary assessment was not made [25]. Using AMSTAR 2 criteria, confidence in the results of 13 out of 17 reviews with included studies was rated as “critically low”, one as “low”, one as “moderate” and two as “high”. Common methodological drawbacks were: omission of prospective protocol submission or publication; use of inappropriate search strategy: lack of independent and dual literature screening and data-extraction (or methodology unclear); absence of explanation for heterogeneity among the studies included; lack of reasons for study exclusion (or rationale unclear).

Risk of bias assessment, based on a clearly reported methodological tool, and quality of evidence appraisal, in line with the Grading of Recommendations Assessment, Development and Evaluation (GRADE) method, were reported only in one review.^16^ Systematic reviews should perform a risk of bias assessment of included primary studies, but only five (27.7%) of the included reviews presented a table summarizing bias. Review authors used a varied number of risk of bias tools. Of these, four reviews classified studies as being of “fair” or “appropriate quality” and one review classified studies as having a high risk of bias. One review mentioned the risk of bias assessment in the methodology but did not provide any related analysis. It is worth mentioning that the reviews used a varied amount of risk of bias tools.

## 4. Discussion

This overview of systematic reviews, to our knowledge the first of its kind on COVID-19, included 18 reviews published up to March 24, 2020, and involving more than 60,000 patients. Ten reviews included meta-analyses. The reviews presented data on clinical manifestations, laboratory and radiological findings and interventions. We found no systematic reviews on the utility of diagnostic tests.

Symptoms were reported in seven reviews; the majority of the patents had fever, cough, dyspnea, myalgia or muscle fatigue and gastrointestinal disorders such as diarrhea, nausea or vomiting. Olfactory dysfunction (anosmia or dysosmia) has been reported in the scientific literature to occur in some patients infected with COVID-19 but this was not reported in any of the assessed systematic reviews [32,33,34]. During the SARS outbreak in 2002, there were reports of impairment of sense of smell associated with the disease [35,36]. Nasal obstruction and post-viral olfactory disorders are possible reasons for this symptom [37].

The reported mortality rates ranged from 0.3 to 14% in the included reviews. Mortality estimates are influenced by the transmissibility rate (basic reproduction number), availability of diagnostic tools, notification policies, asymptomatic presentations of the disease, resources for disease prevention and control and treatment facilities. Even with more than 1,359,398 people infected as of April 7, 2020 [4], variability in the mortality rate fits the pattern of emerging infectious diseases [38]. Furthermore, the reported cases do not consider asymptomatic cases, mild cases where individuals have not sought medical treatment, and the fact that many countries have limited access to diagnostic tests or have implemented testing policies only very recently [39]. The lack of reviews assessing diagnostic testing (sensitivity, specificity and predictive values of RT-PCT or immunoglobulin tests), and the preponderance of studies that assessed only symptomatic individuals, means that considerable imprecision remains around the calculated mortality rate.

Few reviews included treatment data. Those that did described studies offering a very low level of evidence: usually small, retrospective studies with very heterogenous populations. The efficacy of pharmacological interventions therefore remains extremely unclear. Encouragingly, on April 7, 2020 there were 366 clinical trials for COVID-19 registered on ClinicalTrials.gov, with many pharmacological interventions being tested, so hopefully this gap in the evidence will be filled soon [40].

Seven reviews analyzed laboratory parameters. Elevated CRP, lactate dehydrogenase, AST and ALT were commonly reported; elevated cTnI and procalcitonin, and lower platelet count, appear to be associated with more severe disease. This information may be useful for clinicians who attend patients suspected of COVID-19 in emergency services worldwide, for example, to assess which patients need to be reassessed more frequently. Further research is required to assess their relation to mortality and other patient outcomes, as well as prognosis, in order to investigate whether they can contribute to decision-making.

Most systematic reviews scored poorly on the AMSTAR 2 critical appraisal tool for systematic reviews. Thirteen of the 17 included reviews achieved the lowest rating of “critically low” in the confidence readers can have in the results of the review. In contrast, only two reviews achieved the top rating of “high” confidence in the results. Most of the original studies included in the reviews were case series and case reports, which impacts on the quality of evidence. This has major implications for clinical practice and use of these reviews in evidence-based practice and policy. Clinicians, patients and policymakers can only have the highest confidence in systematic review findings if high-quality systematic review methodologies are employed. The urgent need for information during a pandemic does not justify poor quality reporting.

A limitation of our study is that for AMSTAR 2 assessment we relied on information available in publications; we did not attempt to contact study authors for clarifications or additional data. Appraisal of methodological quality of three reviews was hindered because they were published as letters, labelled as rapid communications. As a result, various details about their review process were not included, leading to AMSTAR 2 questions being answered as not reported and thus to low confidence scores. Full manuscripts might have provided additional information that could have led to higher confidence in the results. In other words, low scores could reflect incomplete reporting, not necessarily low quality review methods. In seeking to make their review available more rapidly and more concisely, methodological details may have been omitted. This reflects a general issue during a crisis where speed and completeness must be balanced. Given that some included reviews scored “high”, the two factors are not mutually exclusive. However, this requires proper resourcing and commitment to ensure users of systematic reviews can have high confidence in the results.

Despite this limitation, this overview adds to the current knowledge by providing a comprehensive summary of all the currently available evidence about COVID-19 from published systematic reviews. This overview followed strict methodological criteria, including a comprehensive and sensitive search strategy and a standard tool for methodological appraisal of systematic reviews. Another potential limitation is that several authors from this study have contributed to one of the reviews identified. To reduce risk of any bias, two authors who did not contribute to the review in question undertook the assessment of its quality and limitations.

Further research is of the utmost importance to assess the impact of antivirals, chloroquine and other interventions on patient outcomes. Additionally, it is essential to determine the accuracy of available diagnostic tests. Understanding the role of different biomarkers to predict clinical outcomes, and whether they could be used in association with validated scores, such as Sequential Organ Failure Assessment (SOFA) score, to help the decision of choosing patients for scarce intensive care unit beds would be very useful.

## 5. Conclusion

In conclusion, by conducting this overview of systematic reviews, we have quantified the overall incidence of clinical manifestations of patients infected with SARS-CoV-2 and described its more common laboratory and radiological abnormalities. The evidence is sufficient to guide medical doctors and health-related professionals on the construction of clinical rationale of non-tested cases, but insufficient to provide any specific endorsement of pharmacological and non-pharmacological interventions. In order to fill evidence-based gaps, high quality primary studies should be carried out initially, so that well-designed systematic reviews can be performed in the future.

## Data Availability

All data involved in this submission is fully available.

## Declarations

### Ethics approval and consent to participate

Not required as data was based on primary studies.

### Consent for publication

Not applicable

### Availability of data and materials

All data collected and analyzed within this study are available from the corresponding author on reasonable request.

### Authors’ contributions

IJBN conceived the research idea and worked as a project coordinator. MSM, DPOM, TCVG, HMA, IW, AM, LP, VTC, IZG, TPP and ANA were involved in data curation, formal analysis, investigation, methodology, and initial draft writing. IJBN, MSM, DPOM, SF, NLB and LP analyzed, processed, interpreted the data and wrote the final manuscript draft and also revised it afterwards. All affiliated authors provided feedback to the manuscript, which was coordinated by IJBN and TCVG.

#### Acknowledgements

We thank Catherine Henderson DPhil from Swanscoe Communications for pro bono medical writing and editing support. We acknowledge support from Covidence Team, specifically Anneliese Arno. We thank the whole International Network of Coronavirus Disease 2019 (InterNetCOVID-19) for their commitment and involvement. Members of the InterNetCOVID-19 are listed in Appendix 2. We thank Pavel Cerny and Roger Crosthwaite for guiding the team supervisor (IJBN) on human resources management.

---

This file with figures has been provided by the authors to give readers additional information about their work

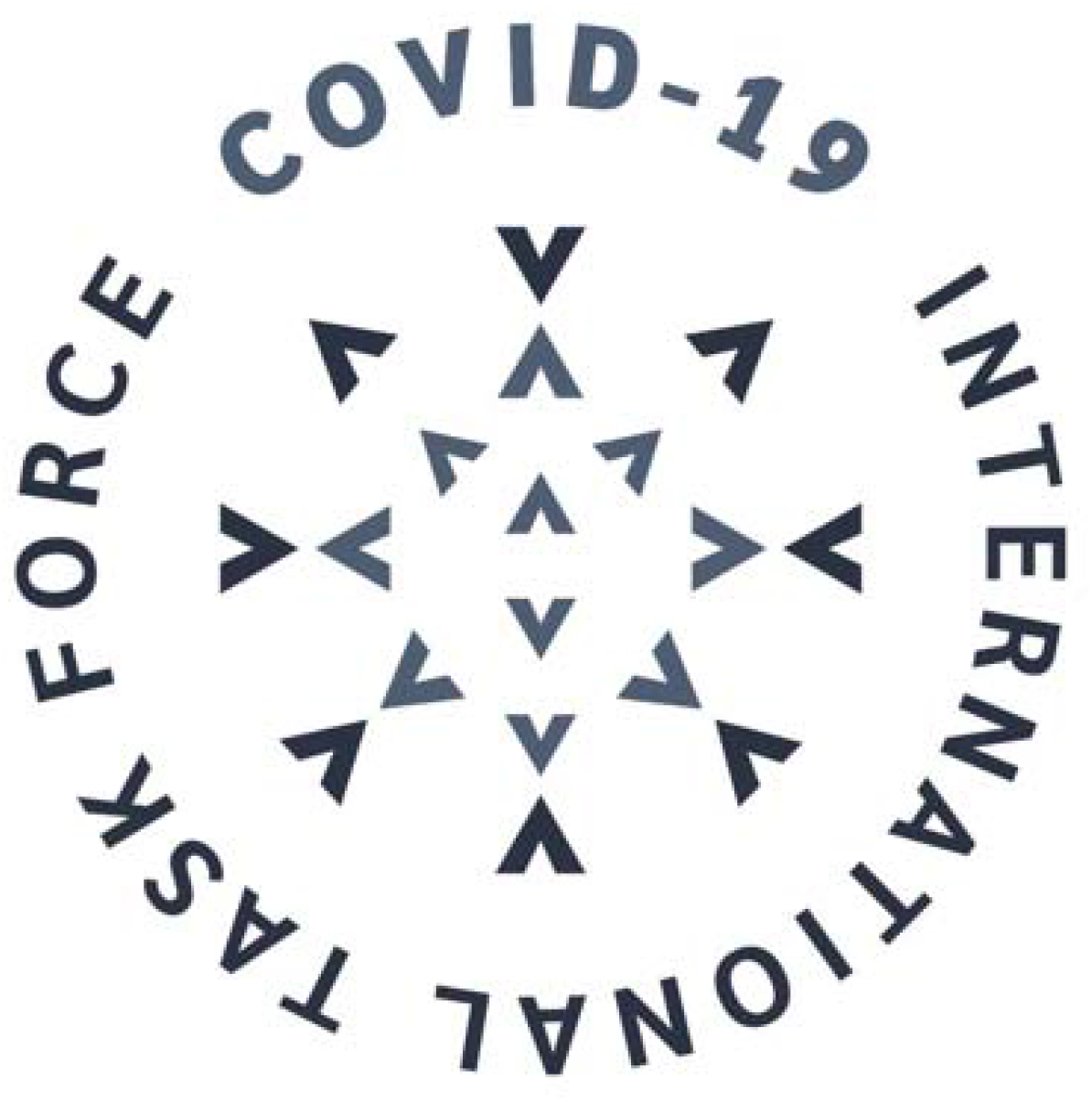

This file with tables has been provided by the authors to give readers additional information about their work

**Table S1.** Descriptive summary of the 18 systematic reviews included in the analysis.

| Review identification | Reference | Journal and IF | Time frame assessed | Databases | Number of included studies | Study design of included studies | Meta-analysis performed | Patients: total (n) | Mean/Median age of included patients (years) | Male sex (%) |
| --- | --- | --- | --- | --- | --- | --- | --- | --- | --- | --- |
| Adhikari et al | <sup>14</sup> | Infectious Diseases of Poverty, 3.12 | January 1, 2020 to January 31, 2020 | BioRxiv, medRxiv, ChemRxiv, Google Scholar, PubMed, CNKI and WanFang Data | 65 | Case reports, case series and observational studies | N | Not reported | Median: 59 | More cases |
| Bo et al | <sup>15</sup> | Clinical Research in Cardiology, 4.907 | December 2019 to February 2020 | Medline, EMBASE | 6 | Case reports, case series and observational studies | Y | 1,527 | Median: 49.75 | 57.80 |
| Borges do Nascimento et al | <sup>16</sup> | Journal of Clinical Medicine, 5.688 | 2019 to February 24, 2020 | MEDLINE, EMBASE, Cochrane Library, Scopus and LILACS | 60 | Case reports, case series and observational studies | Y | 59,254 | Not reported | 51.92 |
| Cortegiani et al | <sup>17</sup> | Journal of Critical Care, 2.783 | Inception up to March 1, 2020 | PubMed and EMBASE, ClinicalTrials.gov, and ICTPR | 6 | Case reports, case series and observational studies | N | 3,090 | Not reported | Not reported |
| Lippi et al <sup>a</sup> | <sup>18</sup> | European Journal of Internal Medicine, 3.660 | 2019 to March 9, 2020 | PubMed, Scopus and Web of Science | 5 | Case reports, case series and observational studies | Y | 1,399 | Not reported | Not reported |
| Lippi et al <sup>b</sup> | <sup>19</sup> | Progress in Cardiovascular Diseases, 6.162 | 2019 to March 4, 2020 | PubMed, Scopus and Web of Science | 4 | Case reports, case series and observational studies | Y | 341 | Not reported | Not reported |
| Lippi et al <sup>c</sup> | <sup>20</sup> | Clinica Chimica Acta, 2.735 | Inception to March 3, 2020 | PubMed, EMBASE, Web of Science | 4 | Case reports, case series and observational studies | Y | Not reported | Not reported | Not reported |
| Lippi et al <sup>d</sup> | <sup>21</sup> | Clinica Chimica Acta, 2.735 | 2019 to March 6, 2020 | PubMed, EMBASE, Web of Science | 9 | Case reports, case series and observational studies | Y | 1,799 | Not reported | 71.71 |
| Li L. et al | <sup>22</sup> | Journal of Medical Virology, 2.049 | December 2019 to February 2020 | PubMed, EMBASE, Wangfang Data and ICTPR | 10 | Case reports, case series and observational studies | Y | 1,994 | Median: 49.94 (without one study) | 63.35 |

| Review identification | Reference | Journal and IF | Time frame assessed | Databases | Number of included studies | Study design of included studies | Meta-analysis performed | Patients: total (n) | Mean/Median age of included patients (years) | Male sex (%) |
| --- | --- | --- | --- | --- | --- | --- | --- | --- | --- | --- |
| Ludvigsson | <sup>23</sup> | Acta Paediatrica, 2.265 | January 1, 2020 to March 18, 2020 | PubMed and EMBASE + ICTPR | 45 | Case reports, case series and observational studies | N | Not summarized | Not reported | Not reported |
| Lupia et al | <sup>24</sup> | Journal of Global Antimicrobial Resistance, 2,469 | November 30, 2019 to February 13, 2020 | PubMed and Cochrane | 13 | Case reports, case series and observational studies |  | 874 | 42.25 (without one study ranging from 19-63) | 53.67 |
| Marasinghe | <sup>25</sup> | Systematic Review, 1.692 | Inception up to February 2020 | PubMed, EMBASE, Google Scholar, and Scopus | 0 | Not applicable | N | Not applicable | - | - |
| Mullins et al | <sup>26</sup> | Ultrasound in Obstetrics & Gynecology, 5.595 | Inception up to March 10, 2020 | PubMed and MedRxiv | 23 | Case reports, case series and observational studies | N | 32 mothers and 30 newborns | Median: 30 | Not applicable |
| Pang et al | <sup>27</sup> | Journal of Clinical Medicine, 5.688 | December 1, 2019 to February 6, 2020 | PubMed, Embase and Cochrane Library | 27 | Case reports, case series and observational studies | N | 656 | Not reported | Not reported |
| Rodriguez-Morales et al | <sup>28</sup> | Travel Medicine and Infectious Disease, 4.868 | January 1, 2020 to February 23, 2020 | PubMed, Scopus and Web of Science | 58 | Case reports, case series and observational studies | Y | 2,874 | Mean: 51.97 | 55.9 |
| Salehi et al | <sup>29</sup> | American Journal of Roentgenology, 3.161 | Inception up to February 19, 2020 | PubMed, EMBASE, Google Scholar, and the World Health Organization database | 30 | Case reports, case series and observational studies | N | 919 | Not reported | Not reported |
| Sun et al | <sup>30</sup> | Journal of Medical Virology, 2.049 | Inception up to February 24, 2020 | PubMed, EMBASE and Cochrane Library | 10 | Case reports, case series and observational studies | Y | 50,466 | Mean: 44.25 | 52.01 |

| Review identification | Reference | Journal and IF | Time frame assessed | Databases | Number of included studies | Study design of included studies | Meta-analysis performed | Patients: total (n) | Mean/Median age of included patients (years) | Male sex (%) |
| --- | --- | --- | --- | --- | --- | --- | --- | --- | --- | --- |
| Yang et al | <sup>31</sup> | International Journal of Infectious Diseases, 3.538 | January 1, 2020 to February 25, 2020 | PubMed, EMBASE and Web of Science | 8 | Case reports, case series and observational studies | Y | 46.248 | Median: 46.0 | 51.6 |

**Table S2.** Main findings observed in the systematic reviews obtained.

| Review identification | Reference | Main findings | Quality of evidence | Overall quality assessment using AMSTAR 2 | Review strengths and limitations |
| --- | --- | --- | --- | --- | --- |

**Table S2. Main findings observed in the systematic reviews obtained**
| Review identification | Reference | Main findings | Quality of evidence | Overall quality assessment using AMSTAR 2 | Review strengths and limitations |
| --- | --- | --- | --- | --- | --- |
| Adhikari et al | 14 | **General summary**<br>The study presents several categories of findings. Epidemiological findings showed that both the immunosuppressed and normal population appear susceptible to the COVID-19 infection. Biological analysis showed that SARS-CoV-2 is similar to $\beta$ coronaviruses found in bats. The effective reproductive number of SARS-CoV-2 (2.9) is higher than that of SARS (1.77). Virus transmission originated in Wuhan and from there it spread internationally. It is likely to be transmitted from human-to-human contact (via droplets or aerosol) as well as via surface contact. Prevention and control consist of isolation, identification and follow-up of contacts, environmental disinfection, and use of personal protective equipment.<br>**Clinical Symptoms**<br>The most commonly reported clinical symptoms are fever, cough, myalgia or fatigue, pneumonia, and complicated dyspnea, whereas less commonly reported symptoms include headache, diarrhea, hemoptysis, runny nose, and phlegm producing cough. Patients with mild symptoms were reported to recover after 1 week while severe cases were reported to experience progressive respiratory failure due to alveolar damage from the virus, which may lead to death.<br>**Diagnosis**<br>Diagnosis of suspected cases used real-time fluorescence (RT-PCR) to detect the positive nucleic acid of SARS-CoV-2 in sputum, throat swabs, and secretions of the lower respiratory tract samples. | Not available | Critically Low | Early scoping report. Broad range of topics addressed.<br>Good review methods, incompletely reported.<br>Narrative presentation of many results. |
| Bo et al | 15 | **Comorbidities evaluated**<br>The most prevalent comorbidities among confirmed cases of COVID-19 were hypertension (17.1%), cardiocerebrovascular disease (16.4%) and diabetes (9.7%). Patients with severe disease/in ICU were more likely to have hypertension, cardio-cerebrovascular diseases and diabetes than patients with non-severe disease/not in ICU; 8.0% of patients with COVID-19 suffered acute cardiac injury. Incidence of myocardial injury was ~13 times higher in patients with severe disease/in ICU than patients with non-severe disease/not in ICU. | Not available | Critically Low | Addresses important comorbidity.<br>Good review methods, incompletely reported. |
| Borges do Nascimento et al | 16 | **General summary**<br>All-cause mortality was 0.3% (95% CI 0.0%–1.0%). Epidemiological studies showed that mortality was higher in males and elderly patients.<br>**Clinical Symptoms**<br>The incidence of symptoms were shown as following: Fever 82%, (CI) 56%–99%; Cough 61%, 95% CI 39%–81%; Muscle aches and/or fatigue 36%, 95% CI 18%–55%; Dyspnea 26%, 95% CI 12%–41%; Headache 12%, 95% CI 4%–23%; Sore throat 10%, 95% CI 5%–17% and gastrointestinal symptoms 9%, 95% CI 3%–17%.<br>**Laboratory findings**<br>Laboratory findings revealed lymphopenia ($0.93 \times 10^9/L$, 95% CI $0.83$–$1.03 \times 10^9/L$ and abnormal C-reactive protein (33.72mg/dL, 95% CI 21.54–45.91mg/dL).<br>**Radiological findings**<br>Radiological findings varied, but mostly described ground-glass opacities and consolidation.<br>**Therapeutic findings**<br>Antivirals (oseltamivir, umifenovir, ganciclovir, ritonavir) were reported as the most commonly used agents. Use of antibiotics was also reported (vancomycin, azithromycin, meropenem, cefaclor, cefepime and tazobactam). Other medications used were corticosteroids, alpha-interferon, immunoglobulin and antifungal drugs. | All-cause mortality with a very low quality of evidence using GRADE | High | Broad, well-conducted review with meta-analysis.<br>Included studies up to 24 February 2020. |

**Table S2. Main findings observed in the systematic reviews obtained**
| Review identification | Reference | Main findings | Quality of evidence | Overall quality assessment using AMSTAR 2 | Review strengths and limitations |
| --- | --- | --- | --- | --- | --- |
| Cortegiani et al | 17 | ***Therapeutic findings***<br>Chloroquine seemed to be effective in limiting the replication of SARS-CoV-2 in vitro, justifying clinical research in patients with COVID-19. However, clinical use should adhere to the Monitored Emergency Use of Unregistered Interventions framework or be ethically approved as a trial. | Not available | Critically Low | Addresses important therapeutic potential. Included reports low on evidence hierarchy. Review methods incompletely reported |
| Lippi et al <sup>a</sup> | 18 | ***Laboratory findings***<br>No significant association was found between active smoking and severity of COVID-19 (OR, 1.69; 95% CI, 0.41–6.92; p=0.254). | Not available | Critically Low | Addresses important comorbidity. Rapid communication (letter) with incomplete reporting. |
| Lippi et al <sup>b</sup> | 19 | ***Laboratory findings***<br>cTnI values were significantly increased in patients with severe COVID-19 compared to those with non-severe disease (SMD, 25.6 ng/L; 95% CI, 6.8–44.5 ng/L). | Not available | Critically Low | Addresses important comorbidity with prognostic value. Rapid communication (letter) with incomplete reporting. |
| Lippi et al <sup>c</sup> | 20 | ***Laboratory findings***<br>Results suggested that an increased procalcitonin value is associated with a higher risk of severe COVID-19 (OR, 4.76; 95% CI, 2.74–8.29). | Not available | Critically Low | Addresses lab value with potential prognostic value. Rapid communication (letter) with incomplete reporting. |
| Lippi et al <sup>d</sup> | 21 | ***Laboratory findings***<br>Platelet count was significantly lower in patients with more severe COVID-19 (weighed mean difference (WMD) $-31 \times 10^9/L$; 95% CI $-35-29 \times 10^9/L$). A subgroup analysis comparing patients by survival, found an even lower platelet count was observed with mortality (WMD, $-48 \times 10^9/L$; 95% CI $-57$ to $-39 \times 10^9/L$). A low platelet count was associated with over fivefold enhanced risk of severe COVID-19 (OR, 5.1; 95% CI 1.8-14.6). | Not available | Critically Low | Addresses lab value with potential prognostic value. Good review methods incompletely reported. |
| Li L. et al | 22 | ***General summary***<br>The patients were 60% male (95% CI [0.54, 0.65]), the discharge rate was 42% (95% CI [0.29, 0.55]), and the fatality rate was 7% (95% CI [0.04,0.10]).<br>***Clinical Symptoms***<br>Clinical symptoms presented were: fever (88.5%), cough (68.6%), myalgia or fatigue (35.8%), expectoration (28.2%), and dyspnea (21.9%), headache or dizziness (12.1%), diarrhea (4.8%), nausea and vomiting (3.9%).<br>***Laboratory findings***<br>Laboratory results showed lymphocytopenia (64.5%), increase of C-reactive protein (44.3%), increase of lactic dehydrogenase (28.3%), and leukocytopenia (29.4%). | Not available | Moderate | Well-conducted review with meta-analysis. Included studies were from China only. |

**Table S2. Main findings observed in the systematic reviews obtained**
| Review identification | Reference | Main findings | Quality of evidence | Overall quality assessment using AMSTAR 2 | Review strengths and limitations |
| --- | --- | --- | --- | --- | --- |
| Ludvigsson | 23 | **General summary**<br>Children account for 1–5% of diagnosed COVID-19 cases and they frequently have milder disease than adults; deaths have been extremely rare. Newborn infants have developed symptomatic COVID-19, but evidence of vertical intrauterine transmission is scarce.<br>**Clinical Symptoms**<br>Clinical characteristics presented mainly as fever and respiratory symptoms, and fewer children seem to have developed severe pneumonia.<br>**Diagnosis**<br>Nasal and pharyngeal swabs or blood analysis are adequate samples for RT-PCR. Sequencing of specimens and clinical diagnosis have been used as alternative diagnostic approaches.<br>**Laboratory findings**<br>Elevated inflammatory markers were less common in children than adults and lymphocytopenia seemed rare.<br>**Radiological findings**<br>Included studies described ground-glass opacities, local or bilateral patchy shadowing, and halo signs.<br>**Therapeutic findings**<br>Suggested treatment included providing oxygen, inhalations, nutritional support and maintaining fluids and electrolyte balances. | Not available | Critically Low | Important population: children.<br>Review methods incompletely reported. Individual studies described narratively without meta-analysis. |
| Lupia et al | 24 | **General summary**<br>Most of the patients were male (age range, 8-92). Cardiovascular, digestive and endocrine system diseases were commonly reported.<br>**Clinical Symptoms**<br>Fever, cough, dyspnea, myalgia and fatigue were the most common symptoms.<br>**Laboratory findings**<br>Case studies reported leucopenia, thrombocytopenia, slightly elevated AST and ALT, and elevated C-reactive protein.<br>**Radiological findings**<br>Multiple bilateral lobular and subsegmental areas of consolidation or bilateral GGOs were most commonly reported in chest CT findings.<br>**Therapeutic findings**<br>Lopinavir, ritonavir, umifenovir and oseltamivir were the most common antivirals used to treat the infection. Supportive intervention (oxygen therapy) was frequently required by patients. Empirical antibiotics have been described. Steroids were also commonly described. | Not available | Critically Low | Summarizes findings from English-language case reports and case series.<br>Review methods incompletely reported. |
| Marasinghe | 25 | No studies were found investigating the effectiveness of face mask use in limiting the spread of this specific virus. |  | NA | Important preventive topic.<br>No data available to review. |

**Table S2. Main findings observed in the systematic reviews obtained**
| Review identification | Reference | Main findings | Quality of evidence | Overall quality assessment using AMSTAR 2 | Review strengths and limitations |
| --- | --- | --- | --- | --- | --- |
| Mullins et al | 26 | **General information**<br>Study revealed that 7 mothers were asymptomatic (21.8%) and 2 mothers were admitted to the intensive care unit (6.25%). Delivery was by Cesarean section in 27 cases and by vaginal delivery in two, and 15 (47%) delivered preterm. There was one stillbirth and one neonatal death. | Not available | Critically Low | Important population: pregnant women. Rapid review methods lowered quality. |
| Pang et al | 27 | **Diagnosis**<br>Possible diagnostic approaches are RT-PCR, serological assays and point-of-care testing.<br>**Therapeutic findings**<br>Several trials were identified, investigating therapeutics such as hydroxychloroquine, lopinavir and ritonavir, glucocorticoids therapy. Several vaccines are in development. | Not available | Critically Low | Review focused on potential new diagnostics and therapeutics. Review methods incompletely reported. |
| Rodriguez-Morales et al | 28 | **General information**<br>20.3% (95% CI 10.0–30.6%) of patients required ICU support, 32.8% presented with acute respiratory distress syndrome (95% CI 13.7–51.8) and 6.2% (95% CI 3.1–9.3) with shock. The case fatality rate was 13.9% (95% CI 6.2–21.5%).<br>**Clinical Symptoms**<br>Clinical symptoms presented were fever (88.7%, 95% CI 84.5–92.9%), cough (57.6%, 95% CI 40.8–74.4%) and dyspnea (45.6%, 95% CI 10.9–80.4%).<br>**Laboratory findings**<br>Regarding laboratory findings, decreased albumin (75.8%, 95% CI 30.5–100.0%), high C-reactive protein (58.3%, 95% CI 21.8–94.7%), and high lactate dehydrogenase (LDH) (57.0%, 95% CI 38.0–76.0), lymphopenia (43.1%, 95% CI 18.9–67.3), and high erythrocyte sedimentation rate (ESR) (41.8%, 95% CI 0.0–92.8), were the most common laboratory results.<br>**Radiological findings**<br>Results showed bilateral pneumonia, with associated ground-glass opacities. | Not available | High quality Review | Broad, well-conducted review with meta-analysis. Included studies up to 23 February 2020. |
| Salehi et al | 29 | **Radiological findings**<br>A correlation was found between CT findings and disease severity and mortality. In severely ill patients, the most commonly reported CT findings were bilateral and multilobar involvement and subsegmental consolidative opacities. ARDS was the most common indication for transfer to the ICU, with the majority of COVID-19 mortalities occurring among patients with ARDS in the ICU. | Not available | Critically Low | Focused review on radiological imaging. Review methods incompletely reported. Only English-language studies included. |
| Sun et al | 30 | **General information**<br>The percentage of severe cases among all infected cases was 0.181 (95% CI: 0.127–0.243), and the case fatality rate was 0.043 (95% CI: 0.027, 0.061).<br>**Clinical Symptoms**<br>Clinical symptom presented were fever 0.891 (95% CI: 0.818–0.945), cough 0.722 (95% CI: 0.657–0.782), muscle soreness or fatigue 0.425 (95% CI: 0.213–0.652). ARDS incidence was 0.148 (95% CI: 0.046–0.296)<br>**Radiological findings**<br>The incidence of abnormal chest computer tomography (CT) was 0.966 (95% CI: 0.921–0.993). | Not available | Low | Broad review of retrospective studies. Review methods incompletely reported. Small number of included studies compared to similar reviews. |

**Table S2. Main findings observed in the systematic reviews obtained**
| Review identification | Reference | Main findings | Quality of evidence | Overall quality assessment using AMSTAR 2 | Review strengths and limitations |
| --- | --- | --- | --- | --- | --- |
| Yang et al | 31 | *Clinical Symptoms*<br>The most common clinical symptoms were fever (91±3%, 95% CI 86–97%), cough (67±7%, 95% CI 59–76%), fatigue (51±0%, 95% CI 34–68%) and dyspnea (30±4%, 95% CI 21–40%).<br>*Comorbidities evaluated*<br>The most common comorbidities were hypertension (17±7, 95% CI 14–22%), diabetes (8±6, 95% CI 6–11%), cardiovascular diseases (5±4, 95% CI 4–7%) and respiratory system diseases (2±0, 95% CI 1–3%). There was a higher likelihood that patients with severe disease had hypertension (OR 2.36, 95% CI: 1.46–3.83), respiratory disease (OR 2.46, 95% CI: 1.76–3.44), or cardiovascular disease (OR 3.42, 95% CI: 1.88–6.22), compared with patients with non-severe disease. | Not available | Critically Low | Important focus on comorbidities. Review methods incompletely reported. |

**Table S3.**
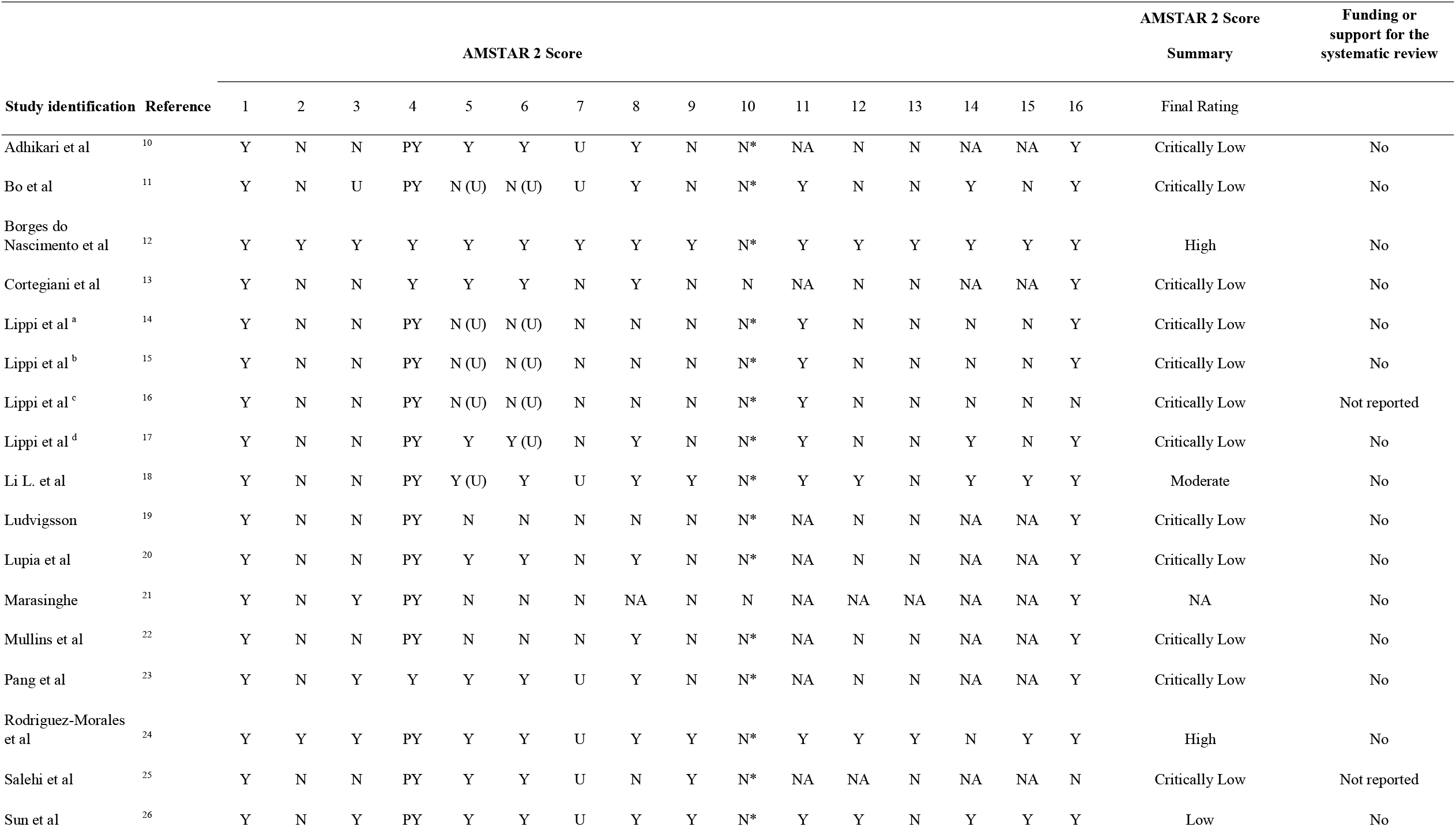
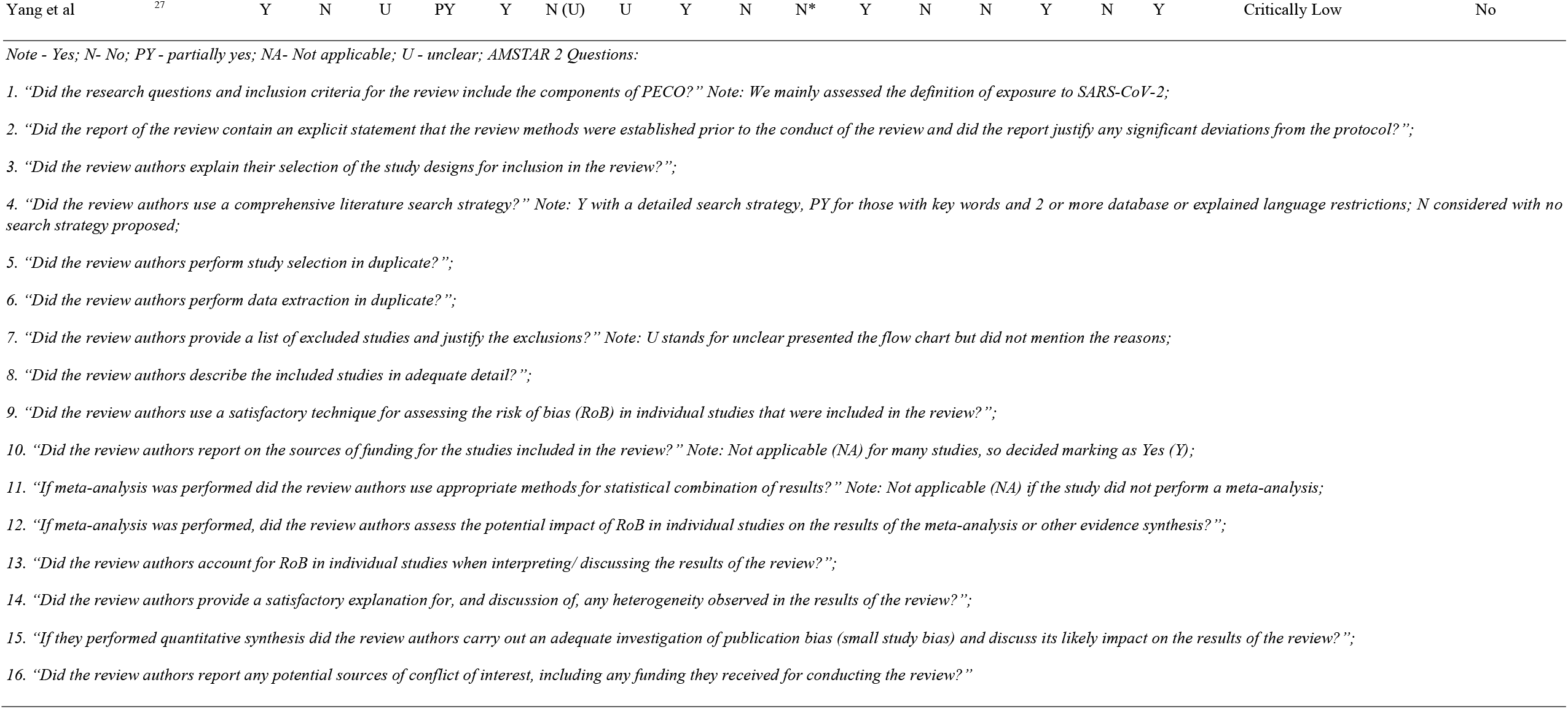
Quality assessment rating of systematic reviews included in the COVID-19 overview.

| Study identification | Reference | AMSTAR 2 Score |  |  |  |  |  |  |  |  |  |  |  |  |  |  |  | AMSTAR 2 Score Summary | Funding or support for the systematic review |
| --- | --- | --- | --- | --- | --- | --- | --- | --- | --- | --- | --- | --- | --- | --- | --- | --- | --- | --- | --- |
|  |  | 1 | 2 | 3 | 4 | 5 | 6 | 7 | 8 | 9 | 10 | 11 | 12 | 13 | 14 | 15 | 16 | Final Rating |  |
| Adhikari et al | <sup>10</sup> | Y | N | N | PY | Y | Y | U | Y | N | N* | NA | N | N | NA | NA | Y | Critically Low | No |
| Bo et al | <sup>11</sup> | Y | N | U | PY | N (U) | N (U) | U | Y | N | N* | Y | N | N | Y | N | Y | Critically Low | No |
| Borges do Nascimento et al | <sup>12</sup> | Y | Y | Y | Y | Y | Y | Y | Y | Y | N* | Y | Y | Y | Y | Y | Y | High | No |
| Cortegiani et al | <sup>13</sup> | Y | N | N | Y | Y | Y | N | Y | N | N | NA | N | N | NA | NA | Y | Critically Low | No |
| Lippi et al <sup>a</sup> | <sup>14</sup> | Y | N | N | PY | N (U) | N (U) | N | N | N | N* | Y | N | N | N | N | Y | Critically Low | No |
| Lippi et al <sup>b</sup> | <sup>15</sup> | Y | N | N | PY | N (U) | N (U) | N | N | N | N* | Y | N | N | N | N | Y | Critically Low | No |
| Lippi et al <sup>c</sup> | <sup>16</sup> | Y | N | N | PY | N (U) | N (U) | N | N | N | N* | Y | N | N | N | N | N | Critically Low | Not reported |
| Lippi et al <sup>d</sup> | <sup>17</sup> | Y | N | N | PY | Y | Y (U) | N | Y | N | N* | Y | N | N | Y | N | Y | Critically Low | No |
| Li L. et al | <sup>18</sup> | Y | N | N | PY | Y (U) | Y | U | Y | Y | N* | Y | Y | N | Y | Y | Y | Moderate | No |
| Ludvigsson | <sup>19</sup> | Y | N | N | PY | N | N | N | N | N | N* | NA | N | N | NA | NA | Y | Critically Low | No |
| Lupia et al | <sup>20</sup> | Y | N | N | PY | Y | Y | N | Y | N | N* | NA | N | N | NA | NA | Y | Critically Low | No |
| Marasinghe | <sup>21</sup> | Y | N | Y | PY | N | N | N | NA | N | N | NA | NA | NA | NA | NA | Y | NA | No |
| Mullins et al | <sup>22</sup> | Y | N | N | PY | N | N | N | Y | N | N* | NA | N | N | NA | NA | Y | Critically Low | No |
| Pang et al | <sup>23</sup> | Y | N | Y | Y | Y | Y | U | Y | N | N* | NA | N | N | NA | NA | Y | Critically Low | No |
| Rodriguez-Morales et al | <sup>24</sup> | Y | Y | Y | PY | Y | Y | U | Y | Y | N* | Y | Y | Y | N | Y | Y | High | No |
| Salehi et al | <sup>25</sup> | Y | N | N | PY | Y | Y | U | N | Y | N* | NA | NA | N | NA | NA | N | Critically Low | Not reported |
| Sun et al | <sup>26</sup> | Y | N | Y | PY | Y | Y | U | Y | Y | N* | Y | Y | N | Y | Y | Y | Low | No |
| Yang et al | 27 | Y | N | U | PY | Y | N (U) | U | Y | N | N* | Y | N | N | Y | N | Y | Critically Low | No |

*Note - Yes; N- No; PY - partially yes; NA- Not applicable; U - unclear; AMSTAR 2 Questions:*

